# The CCR5-delta32 variant might explain part of the association between COVID-19 and the chemokine-receptor gene cluster

**DOI:** 10.1101/2020.11.02.20224659

**Authors:** Juan Gómez, Elías Cuesta-Llavona, Guillermo M. Albaiceta, Marta García-Clemente, Carlos López-Larrea, Laura Amado-Rodríguez, Inés López-Alonso, Tamara Hermida, Ana I. EnrÍquez, Helena Gil, Belén Alonso, Sara Iglesias, Beatriz Suarez-Alvarez, Victoria Alvarez, Eliecer Coto

**Affiliations:** Genética Molecular, Hospital Universitario Central Asturias, Oviedo, Spain; Unidad de Cuidados Intensivos Cardiológicos, Hospital Universitario Central Asturias, Oviedo, Spain; Neumología, Hospital Universitario Central Asturias, Oviedo, Spain; Inmunología, Hospital Universitario Central Asturias, Oviedo, Spain; Urgencias, Hospital Universitario Central Asturias, Oviedo, Spain; Instituto de Investigación Sanitaria del Principado deAsturias, ISPA, Oviedo, Spain; Universidad de Oviedo, Oviedo, Spain; CIBER-Enfermedades Respiratorias. Instituto de Salud Carlos III. Madrid, Spain; Instituto Universitario de Oncología del Principado de Asturias. Oviedo, Spain; Red de Investigación Renal (REDINREN), Madrid, Spain

**Keywords:** COVID-19, SARS-Cov-2, CCR5 delta32, genetic susceptibility

## Abstract

A polymorphism in the *LZTFL1* gene located in the chemokine-receptor gene cluster (chromosome 3p) has been associated with the risk of developing COVID-19. The chemokine receptor-5 (CCR5) maps to this region, and the common 32 bp deletion variant (Δ32) has been associated with the extent of inflammatory disease and the outcome in several viral diseases. Several studies have also suggested that the pharmacological targeting of CCR5 could reduce the impact of SARS-CoV-2 infection and the severity of COVID-19. We sought to investigate whether this polymorphism was associated with the risk of moderate-severe COVID-19.

We genotyped 294 patients who required hospitalization due to COVID-19 (85 were severe cases) and 460 controls. We found a significantly lower frequency of CCR5-Δ32 among the COVID-19 patients (0.10 vs 0.18 in controls; p=0.002, OR=0.48, 95%CI=0.29-0.76). The difference was mainly due to the reduced frequency of CCR5-Δ32 carriers in the severe, significantly lower than in the non-severe patients (p=0.036). Of note, we did not find deletion-homozygotes among the patients compared to 1% among controls. We also confirmed the association between a *LZTFL1* variant and COVID-19. Our study points to CCR5 as a promising target for treatment of COVID-19, but requires validation in additional large cohorts. In confirmed by others, the genetic analysis of CCR5-variants (such as Δ32) might help to identify patients with a higher susceptibility to severe COVID-19.

## Introduction

The clinical spectrum of COVID-19 varies from none or very mild-symptoms to moderate symptoms that required hospitalization, and in some cases severe pneumonia and acute respiratory distress syndrome (ARDS) that requires mechanical ventilation and admission into intensive care unit (ICU). An exacerbated immune response (*cytokine storm*) drives the severity of COVID-19 (**Bohn et al., 2020; Tang et al., 2020; Xiong et al., 2020**). Among other findings, COVID-19 patients showed elevated levels of IL-6 and other proinflammatory cytokines, chemokines, and chemokine-receptors (**Han et al., 2020; Wang et al., 2020; Zeng et al., 2020**).

The C-C chemokine-receptor-5 (CCR5) is expressed on several cell types, including T cells, macrophages and dendritic cells. CCR5 binds to the chemokine RANTES/CCL-5, that has been reported to be increased in COVID-19. CCR5 might thus play a role in inflammatory responses to coronaviruses infection. Human dendritic cells infected by SARS-CoV-1 showed increased CCR5 expression (**Law et al., 2009**). A single-cell RNA sequencing on nasopharyngeal and bronchial samples from patients with moderate or critical disease and healthy controls pointed to CCR5 pathways as suppressors of immune hyperactivation in critical COVID-19 (**Chua et al., 2020**). The levels of ccr5 mRNA were increased in mice infected with the SARS-Cov (**Chen et al., 2010**). It has been reported that mice deficient in CCR5 infected with SARS-CoV-1 exhibited defects in directing inflammatory to the airway compared to wild-type mice (**Sheahan et al., 2008**). Mice lacking the CCR5 infected with a neurotropic coronavirus showed reduced macrophage infiltration and demyelination (**Glass et al., 2001**).

A recent study showed a beneficial effect of leronlimab (a CCR5 blocking antibody) on a reduced group of severe COVID-19 patients (**Patterson et al., 2020**). These patients showed a significant reduction of the SARS-Cov-2 viral load, with a rapid reduction of plasma IL-6 and restoration of the CD4/CD8 ratio. In another study a total of 23 hospitalized severe/critical COVID-19 patients received leronlimab subcutaneously (**Yang et al., 2020**). By day 30 after initial dosing Leronlimab was safe and well tolerated, 17/23 were recovered and 4/23 had died. The authors concluded that those with lower inflammatory markers had better outcomes, with IL-6 tending to fall overtime. Cenicriviroc (CVC) is a CCR5 antagonist with potent and selective anti-human immunodeficiency virus type 1 (HIV-1) activity. CVC showed anti SARS-Cov-2 activity by inhibition of virus-induced cell destruction and reduction of the viral RNA levels in culture supernatants of infected cells (**Okamoto et al., 2020**).

CCR5 is the receptor for HIV entry into the host cells, and a common variant that abrogates its expression in homozygotes (the 32 bp deletion, Δ32) is associated with the resistance to AIDS among HIV-exposed (**Kou and Kuang, 2019**). Approximately 1% of Europeans are Δ32-homozygotes (**Figure 1**), and carriers of this variant would have a reduced inflammatory response. This effect on CCR5-mediated responses could explain the association of Δ32 with several immune-mediated diseases (**Hutter and Ganepola, 2011; González et al., 2001; Soto-Sánchez et al., 2010**). CCR5-Δ32 might also influence the immunological response to several viruses (**Ellwanger et al., 2020)**. The first COVID-19 Genome Wide Association study (GWAs) reported a strong association with chromosome 3 variants in the chemokine-receptor cluster (**Ellinghaus et al., 2020**). CCR5 maps to this region and could thus partly explain the observed association with polymorphisms in the chemokine-receptor cluster. We sought to investigate whether CCR5-Δ32 was associated with the risk of developing COVID-19 or was a genetic modifier of disease severity.

**Figure 1.**
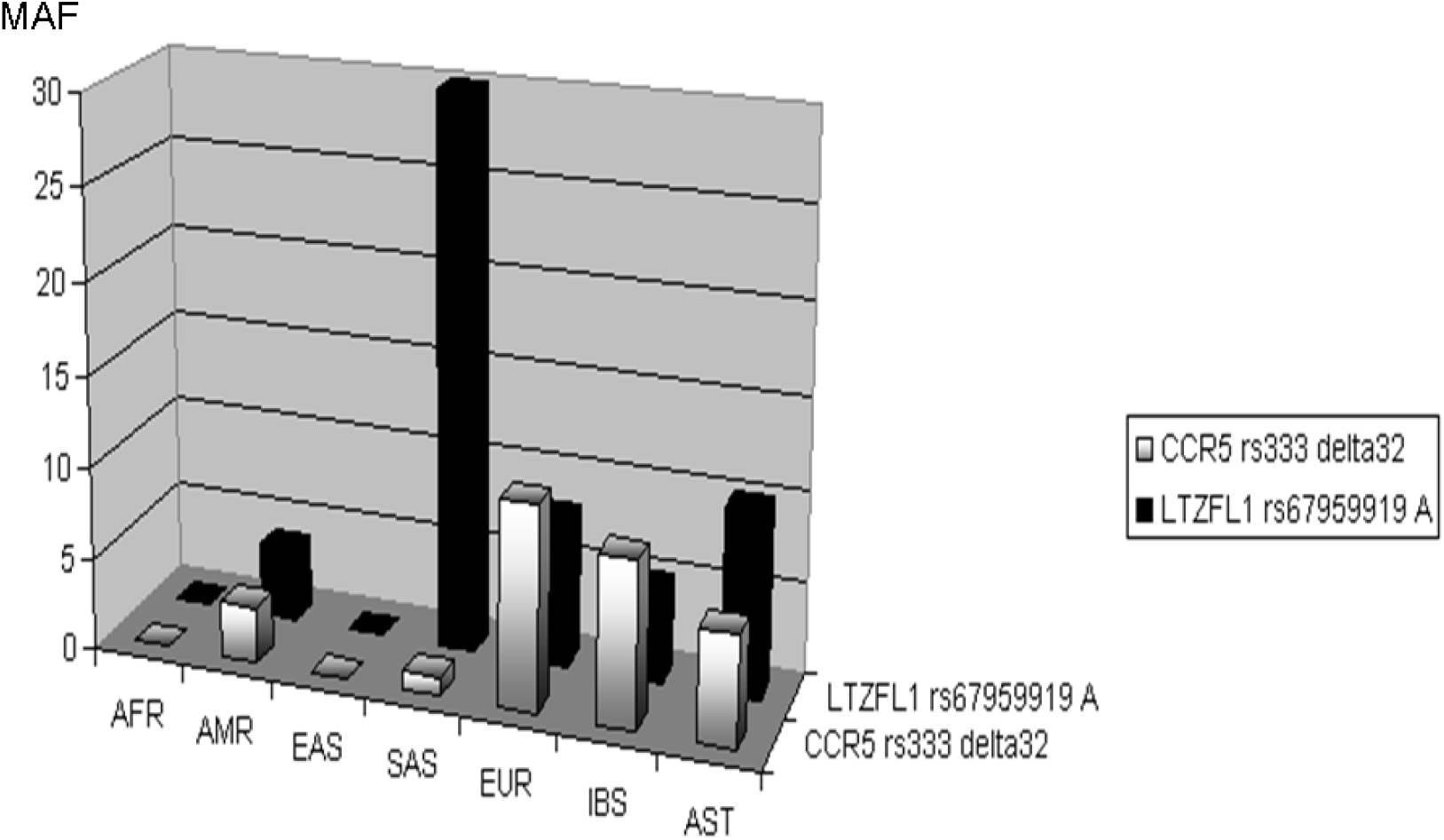
Minor allele frequency of the *CCR5* and *LTZFL1* variants in different populations. Africans (AFR), Americans (AMR), East Asians (EAS), South Asians (SAS), Europeans (EUR), Iberians from Spain (IBS), and Asturians (AST, the study population from Spain). Data obtained from https://www.ensembl.org.

## Patients and Methods

The study was based on 294 patients who required hospitalization due to COVID-19 (mean age 64.77 years, range 24-95) and 460 population age-matched controls (mean age 70.01 years, range 50-81). All the study participants were Caucasian from the region of Asturias (Northern Spain, total population 1 million). Severe cases (n=85) were defined as those in need of critical care support, including high-flow oxygen, positive-pressure ventilation (either invasive or non-invasive) or vasoactive drugs. The presence of comorbidities (hypertension, diabetes, hypercholesterolaemia) was obtained from the participants medical records. The study was approved by the Ethics Committee of Principado de Asturias (Oviedo, Spain), and all the patients or their representatives gave their consent to participate.

We genotyped two variants closely linked in the chromosome 3 chemokine receptor cluster, *LZTFL1* rs67959919 and *CCR5*-Δ32 (rs333). The genomic DNA was PCR-amplyfied with primers flanking the two polymorphic sites (**supplementary figure**). SNP rs67959919 was in complete linkage disequilibrium with rs11385942, associated with COVID-19 in a recent GWAs (**supplementary figure**) (**Ellinghaus et al., 2020**). For this SNP the PCRs were digested with the restriction enzyme *MspI* and the allele-fragments visualised by agarose gel electrophoresis (**supplementary figure**). For the Δ32 the PCRs were directly electrophoresed on 4% agarose gels (**González et al., 2001; Soto-Sánchez et al., 2010**). The accuracy of the genotyping methods was validated by sequencing samples representative of the genotypes (**figure 2**). All the participant′s data (age, sex, COVID-19 severity, hypertension, and genotypes) were collected in an excel file and in accordance to the requirements of the Ethical Committee. Statistical analysis was performed with the R-project free software (www.r-project.org). Logistic regression (linear generalized model, LGM) was used to compare mean values and frequencies between the groups.

**Figure 2.**
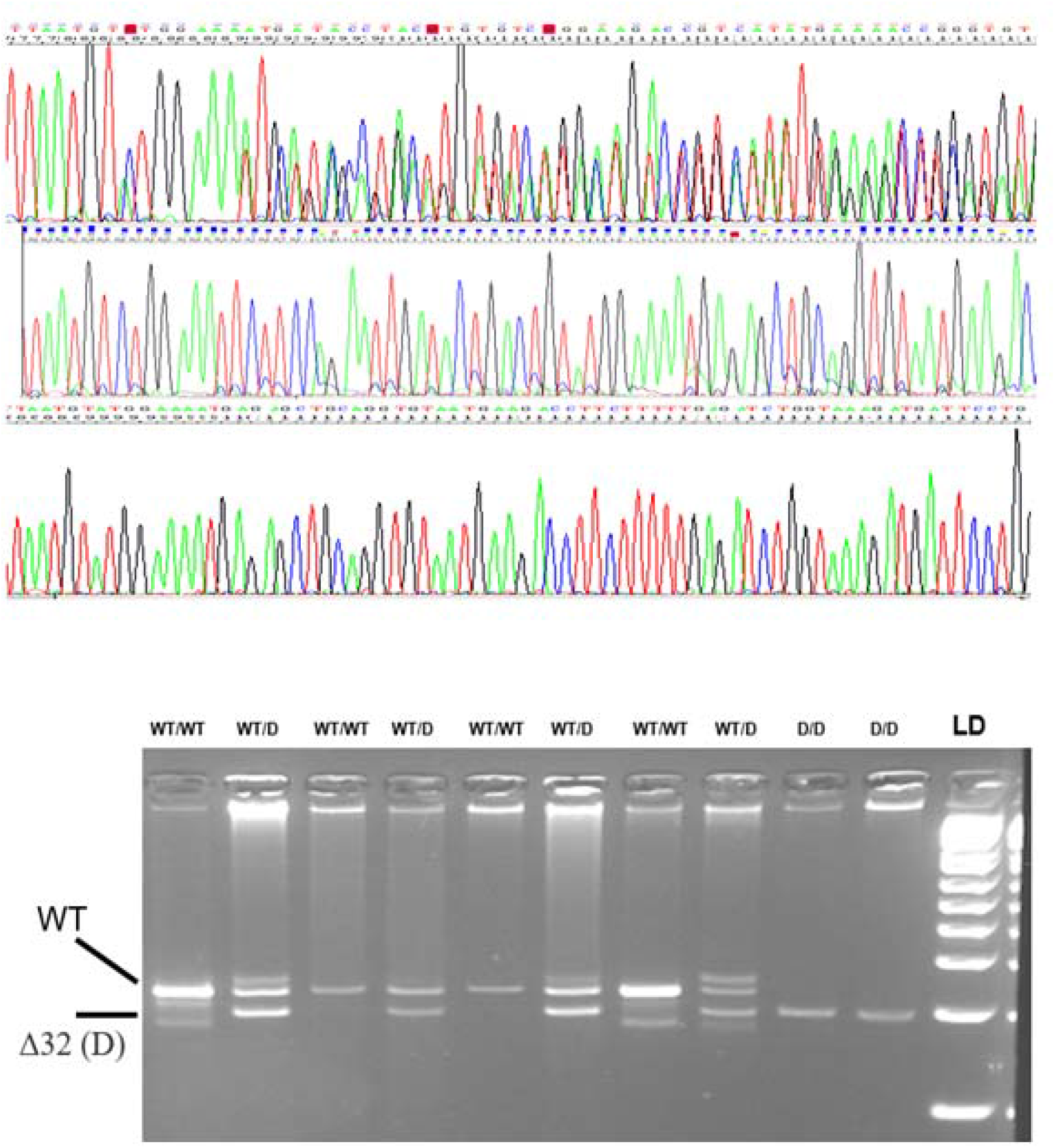
Sequence electropherograms (above) and gel electrophoresis of PCR fragments (below) corresponding at the three CCR5 32 bp deletion. The DNA was amplified with CTGTGTTTGCGTCTCTCCCA and CCTCTTCTTCTCATTTCGACAC, and the PCRs electrophoresed on a 4% agarose gel to visualise the alleles as fragments of 222 bp (wild-type) and 190 bp (Δ32).

## Results and discussion

The main finding of our study was a significantly lower frequency of CCR5-Δ32 among the COVID-19 patients (**table 1**). Multiple logistic regression with age, sex and hypertension showed that in our population carriers of the 32 bp deletion were significantly less frequent among the patients (30/294=0.10 vs 85/460=0.18 in controls; p=0.002, OR=0.48, 95%CI=0.29-0.76). The difference was mainly due to the reduced frequency of CCR5-Δ32 carriers in the severe-ICU group, significantly lower than in the non-severe patients (p=0.036). Of note, we did not find deletion-homozygotes among the patients compared to 1% among controls. Carriers of the deletion would express less membrane-bound receptor and an attenuated pro-inflammatory response mediated by the ligand binding to CCR5. This might explain the reported protective effect of Δ32 on the development of clinical manifestations with underlying inflammatory mechanisms, such as atherosclerosis and coronary heart disease (**Muntinghe et al., 2009; González et al., 2001; Vangelista and Vento, 2018)**. In addition to the well-known protective effect against HIV-1, this variant has been investigated in reference to the clinical outcomes of several viral infections, such as hepatitis B and influenza A **(Ruiz-Mateos et al., 2018; Ellwanger et al., 2020)**. Because CCR5 is not a recognised receptor for SARS-Cov-2 the most likely explanation for the protective effect of Δ32 in COVID-19 is an attenuated inflammatory response among the CCR5-deletion carriers. This finding was in agreement with a report that showed that mice lacking CCR5 infected with a neurotropic coronavirus showed an attenuated inflammatory response, with reduced macrophage infiltration and demyelination (**Glass et al., 2001**).

**Table 1.** Main values in the COVID19 patients and controls. Multiple logistic regression p-values, Odds ratios (OR) and 95% confidence intervals (CI) were calculated including age, sex, hypertension and the corresponding genotypes. IQ=interquartile

|  | COVID<br>N=294 | CONTROLS<br>N=460 | SEVERE<br>N=85 | NON-SEVERE<br>N=209 |
| --- | --- | --- | --- | --- |
| Male | 176 (60%) | 250 (54%) | 64 (75%) | 112 (55%) |
| Mean age, SD, years | 63.18±14.75 | 70.48±6.94 | 65.74±11.16 | 62.14±15.89 |
| Age IQ range, years | 53-75 | 65-76 | 50-75 | 60-75 |
| Hypertension | 131 (45%) | 154 (34%) | 49 (58%) | 82 (39%) |
| ***CCR5* Δ32bp**<br>**rs333** |  |  |  |  |
| Δ / Δ | 0 | 4 (1%) | 0 | 0 |
| Δ / WT | 30 (10%) | 81 (18%) | 4 (6%) | 26 (14%) |
| WT / WT | 264 (90%) | 375 (81%) | 81 (95%) | 183 (88%) |
| Δ32 allele | 0.05 | 0.06 | 0.05 | 0.10 |
| p, OR (95%CI)* | **P=0.002, OR=0.48 (0.29-0.76)** |  | **P=0.036, OR=0.30 (0.09-0.84)** |  |
| ***LZTFL1***<br>**rs67959919** |  |  |  |  |
| AA | 3 (1%) | 4 (1%) | 0 | 3 (1%) |
| AG | 59 (20%) | 60 (13%) | 22 (26%) | 37 (18%) |
| GG | 232 (79%) | 396 (86%) | 63 (74%) | 169 (81%) |
| A allele | 0.13 | 0.10 | 0.11 | 0.07 |
| p, OR (95%CI)** | **P=0.003, OR=1.86 (1.23-2.82)** |  | **P=0.13, OR=1.62 (0.86-3.02)** |  |

\* Δ32 – carriers vs. WT/WT
\*\* A-carriers vs GG

We confirmed the association between ***LZTFL1 (***SNP rs67959919) and the risk of COVID-19. ***LZTFL1*** variants were associated with COVID-19 in a GWAs including participants from Italy and Spain. In our cohort the rare T-allele was associated with an increased risk of COVID-19 after correcting by age, sex and hypertension (**P=0**.**003, OR=1**.**86 (1**.**23-2**.**82)**, but we did not find significant difference between severe and non-severe cases (**Table 1**). We calculated the Δ32 - rs67959919 haplotype frequencies in patients and controls (**table 2**). The rs67959919 A – CCR5 non-deletion (wild-type) haplotype was significantly more common among the patients (p<0.001, OR=2.01, 95%CI=1.45-2.79). We cannot exclude that the association between one of the variants was a consequence to its linkage-disequilibrium (LD) with the other, or with some not yet identified variant in the chemokine-receptor cluster. Multiple logistic regression with the two genotypes were independent predictors of COVID-19: *CCR5*-Δ32 carriers, p=0.005, OR=0.52 (95%CI=0.33-0.81); rs67959919-A carriers, p=0.024, OR=1.56 (95%CI=1.06-2.31).

**Table 2.** Haplotype frequencies in controls, COVID19, and Europeans (EURX, obtained from https://ldlink.nci.nih.gov/?tab=ldpair). The two markers were linked with D′=1.0, r^2^ =0.015. The rs67959919 A – *CCR5*-WT haplotype was significantly more common among the patients (p<0.001, OR=2.01, 95%CI=1.45-2.79). The rs67959919 G – *CCR5*-Δ32 haplotype was significantly less common in the patients (p<0.001, OR=0.25, 95%CI=0.17-0.37).

| **rs67959919 – rs333** | **CONTROLS** | **COVID-19** | **EURX** |
| --- | --- | --- | --- |
| **A – Δ32** | **0** | **0** | **0** |
| **A - WT** | **114 (6%)** | **61 (11%)** | **16 (8%)** |
| **G - Δ32** | **177 (10%)** | **30 (6%)** | **21 (11%)** |
| **G -WT** | **1,554 (84%)** | **429 (83%)** | **161 (81%)** |

The main limitation of our study was the reduced size of the patient′s cohort. However, our results were in agreement with the reported association between chromosome 3p21.31 variants and COVID-19. In this way, the association between *CCR5*-Δ32 and COVID-19 seems plausible and deserves further validation in larger cohorts. If confirmed, the *CCR5* variants might serve as valuable markers to identify individuals predisposed to severe COVID-19. A limitation was also that we did not provide a functional link between the CCR5 deletion and COVID-19 outcomes, and we cannot conclude whether this variant was associated with an overall resistance to SARS-Cov-2 infection or a reduced inflammatory response, or both. Although the CCR5 has not a recognised role in SARS-CoV-2 binding to the host cell, a preliminary study showed that the CCR5 inhibitor Maraviroc was able to decrease the extent of viral-cell fusion and the viral load (**Risner** et al. 2020). To determine whether CCR5 has a direct role in SARS-CoV-2 infection studies with mice expressing the human viral receptor (ACE-2) and lacking ccr5 should be of upmost relevance (**Winckler et al., 2020; Israelow et al., 2020**).

Finally, the pharmacological blockade of *CCR5* has been associated with an improvement of the symptoms in a group of severe COVID-19 patients. In a limited study involving 10 patients the anti-CCR5 antibody Leronlimab showed an improvement of disease symptoms, with a rapid reduction of plasma IL-6 and restoration of the CD4/CD8 ratio and a significant reduction of the SARS-Cov-2 viral load (**Patterson et al., 2020**). Also, in-vitro studies with SARS-Cov-2 infected cells showed that the CCR5 antagonist Cenicriviroc had potent anti-viral activity by inhibition of virus-induced cell destruction and reduction of the viral RNA levels (**Okamoto et al., 2020**). These and other studies pointed to CCR5 as a potential target for COVID-19 treatment. The genetic analysis of CCR5-variants, such as Δ32, might help to identify patients with a higher susceptibility to severe COVID-19.

## Data Availability

data are available upon requesting to the corresponding author at

## Contributorship

All the authors contributed to this work by recruiting the patients and performing the genetic and statistical analysis. E.C. wrote the ms. and takes full responsibility for the accuracy of the data. All the authors approved the submission of this ms.

## Data accessibility

To facilitate the revision of our results by other researchers, an excel file with the data is provided as supplementary file.

## Acknowledgements

This work was supported by a grant from the Spanish Plan Nacional de I+D+I Ministerio de Economía y Competitividad and the European FEDER, grant ISCIII-Red de Investigación Renal-REDINREN RD16/9/5 (EC).

