## SUPPLEMENTAL FILES for "The CCR5-delta32 variant might explain part of the association between COVID-19 and the chemokine-receptor gene cluster"

|  | **SEVERE-ICU, N=85** | | | |
| --- | --- | --- | --- | --- |
| **CCR5**  **D32** | ***LZTFL1*** rs67959919 | | | |
|  | **AA** | **AG** | **GG** | **N** |
| **DD** | **0** | **0** | **0** | **0** |
| **WT/D** | **0** | **0** | **4** | **4** |
| **WT/WT** | **0** | **22** | **59** | **81** |
| **N** | **0** | **22** | **63** |  |
|  | **NON SEVERE, N=209** | | | |
| **DD** | **0** | **0** | **0** | **0** |
| **WT/D** | **0** | **3** | **23** | **26** |
| **WT/WT** | **3** | **34** | **146** | **183** |
| **N** | **3** | **37** | **169** |  |
|  | **CONTROLS, N=460** | | | |
| **DD** | **0** | **0** | **4** | **4** |
| **WT/D** | **0** | **6** | **75** | **81** |
| **WT/WT** | **4** | **54** | **317** | **375** |
| **N** | **4** | **60** | **396** |  |

**Supplementary table.** Number of individuals with each CCR5- *LZTFL1* genotype pairs. Haplotype frequencies were derived from these values.

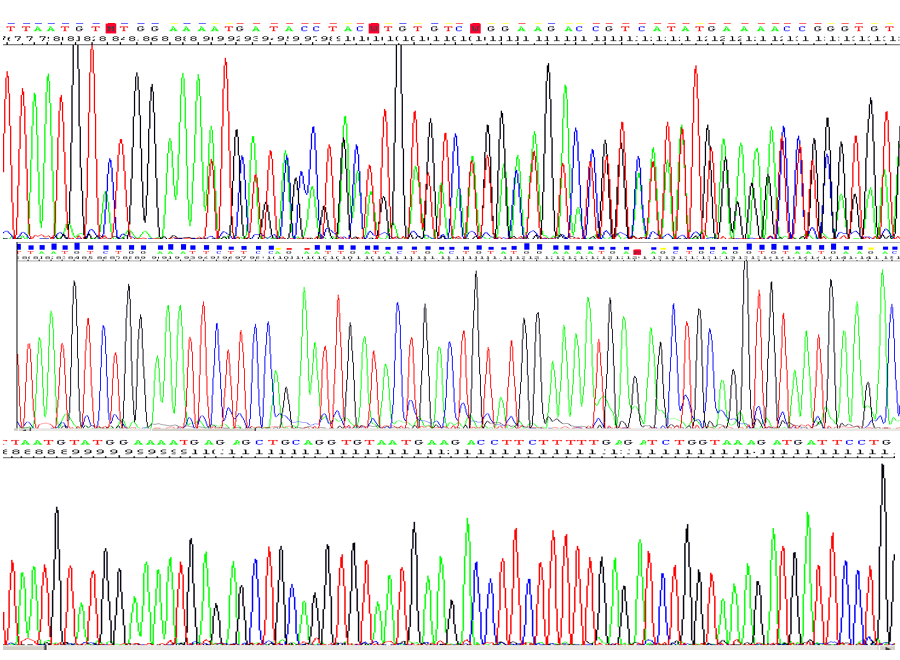

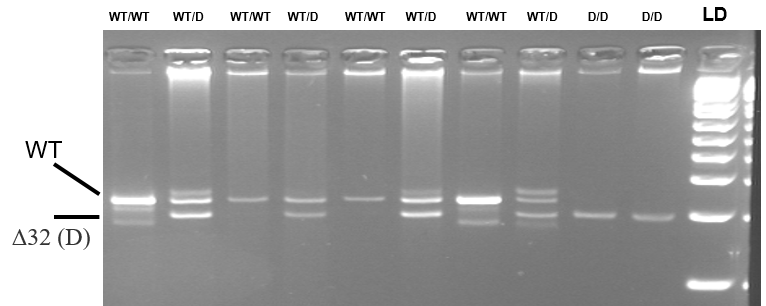

**Supplementary figure.** Sequence electropherograms (above) and gel electrophoresis of PCR fragments (below) corresponding at the three CCR5 32 bp deletion. The DNA was amplified with CTGTGTTTGCGTCTCTCCCA and CCTCTTCTTCTCATTTCGACAC, and the PCRs electrophoresed on a 4% agarose gel to visualise the alleles as fragments of 222 bp (wild-type) and 190 bp (Δ32).

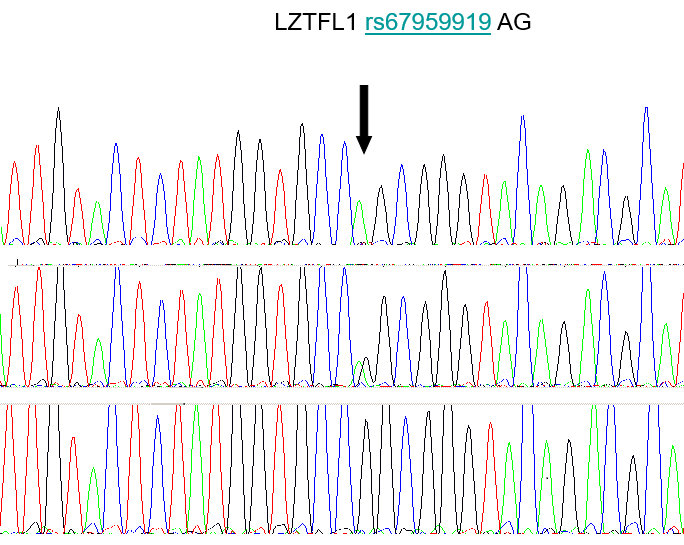

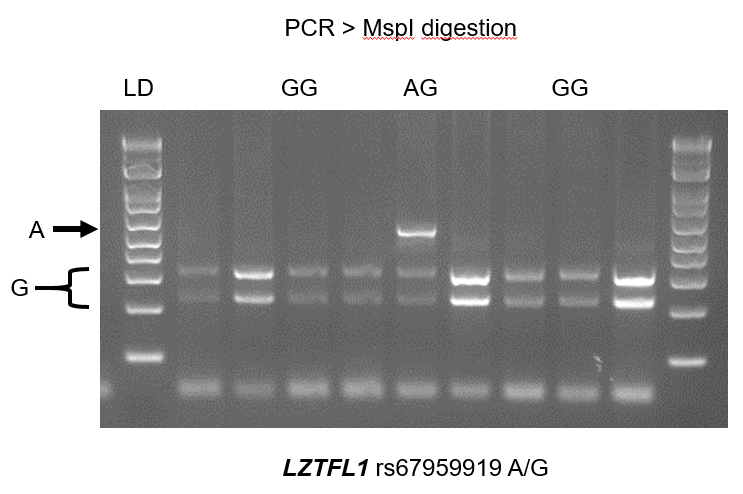

**Supplementary figure.** Sequence electropherograms (above) and gel electrophoresis of PCR fragments (below) corresponding at the three *LZTFL1* genotypes. The DNA was amplified with primers 5´TCCCTCTGTCCATCCTCTAGGGC and 5´GCAATGAGA GTATGACCACTAGAAAAGCC, and digested with MspI. Digestions were electrophoresed on a 4% agarose gel to visualise the two alleles.

LD: 1 kb ladder, DNA size marker.

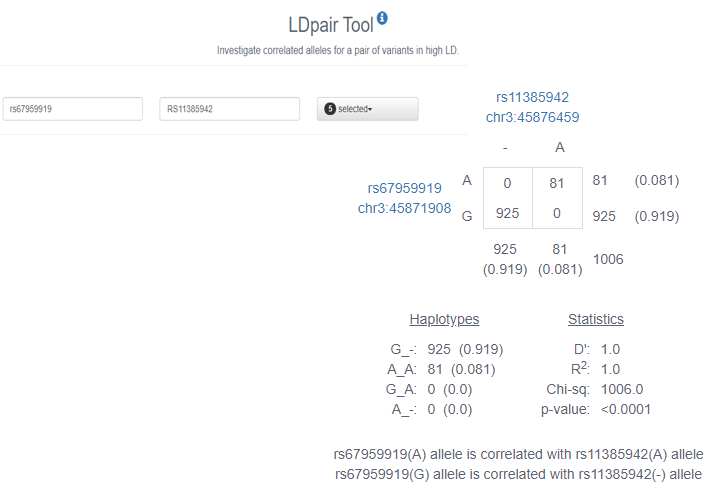

**Suppl. figure.** Haplotype frequencies between the *LZTFL1* rs67959919 and rs11385942 SNPs. The latter was associated with the risk of COVID-19 I a GWAs with Spanish and Italian patients and controls. Data corresponded to Europeans, accessed at the https://ldlink.nci.nih.gov/?tab=ldpair.
